# The Investigation of Nonlinear Variability Underlying Postural Control in The Injure-Limb in Individuals with and without Chronic Ankle Instability

**DOI:** 10.1101/2024.03.11.24304094

**Authors:** Yuki A. Sugimoto, Patrick O. McKeon, Christopher K. Rhea, Randy J. Schmitz, Robert A. Henson, Carl G. Mattacola, Scott E. Ross

**Affiliations:** Department of Physical Therapy and Human Movement Sciences, Northwestern University Feinberg School of Medicine, Chicago, IL 60611; Department of Kinesiology, The University of North Carolina at Greensboro, Greensboro, NC 27402; Department of Exercise Science and Athletic Training, Ithaca College, Ithaca, NY 14850; College of Health Sciences, Old Dominion University, Norfolk, VA, USA

**Keywords:** Nonlinear Dynamics, Sensory Reweighting System, Postural Control, Ankle Sprains

## Abstract

**Background:** Less flexible and adaptable sensorimotor systems reflect in movement variability in individuals with Chronic Ankle Instability (CAI), which may limit their ability to detect relevant information using a variety of primary sensory feedback. Thus, the aim of the study was to investigate underlying biological noise pertaining to postural control in single-limb stance during increased environmental constraints with sensory feedback manipulations in individuals with and without CAI.

**Methods:** Forty-two individuals with and without CAI participated in the study. A one-way ANOVA was utilized to examine group differences in biological noise underlying postural control during the SOT conditions in single-limb stance.

**Results:** Individuals with CAI demonstrated significantly lower SampEN while maintaining posture during Condition 5 (*P*=.037) and Condition 6 (*P*=.030), where they were forced to exclusively rely on vestibular feedback, in single-limb stance compared to healthy controls.

**Discussion:** Individuals with CAI did not demonstrate decreased movement variability pertaining to postural control during all six SOT conditions. Those participants with CAI only displayed decreased movement variability when they were forced to executively rely on vestibular feedback while maintaining posture in the injured-limb compared to healthy controls.

## Introduction

Ankle sprains affect up to two million people in the United States each year, making them one of the most prevalent musculoskeletal injuries among the general public and athletes of all levels who participate in physical activity and athletics [1]. More than 628,000 lateral ankle sprains (LAS) are treated in the emergency room each year, and LAS account for roughly 92% of doctor office visits for ankle sprains [2]–[4]. The vast majority of individuals who have had an initial LAS develop chronic ankle instability (CAI), which is characterized by recurrent episodes of ankle instability [5], [6]. Furthermore, CAI has a negative effect on the quality of life, a decrease in physical activity, and the emergence of long-term deficiencies in activities of daily living [7]. The resulting abnormalities in CAI patients frequently manifest as postural control dysfunction and maladapted gait.

Many postural control studies have looked at the area, velocity, and excursion length of the center-of-pressure (COP) in individuals with CAI. However, there is conflicting evidence showing that individuals with CAI have impaired postural control in unilateral stance when compared to healthy controls [8]–[13]. The lack of consistency could be attributed to the fact that standard COP measures do not specify the temporal proximity to stability boundaries that individuals should strive for when maintaining an upright posture [14]. In contrast, it has been suggested that a nonlinear method known as time-to-boundary and sample entropy (SampEN) could be useful in identifying postural control impairments associated with CAI and other pathological conditions [15]–[17]. SampEN is intended to quantify the amount of regularity of fluctuations in time-series data and detects subtle physiological changes (i.e., neural control) in response to ever-changing environments (environmental constraints). As a result, a nonlinear analysis of SampEN is proposed to quantify the flexibility and adaptability of sensorimotor systems by reflecting biological noise underlying postural control [18]. Hence, incorporating SampEN as a dependent variable can capture the complex nature of the human body with multiple interactions between internal and external properties that traditional COP measures previously overlooked.

Individuals must be able to perceive relevant information associated with the environment using various combinations of somatosensory, visual, and vestibular feedback to achieve optimal performance in unpredictable, ever-changing situations. An initial ankle sprain causes somatosensory dysfunction, and individuals with CAI exhibit less flexible and adaptable sensorimotor systems [19]. The physiological characteristics of CAI may cause individual perception to diversify or isolate sensory systems. Indeed, when compared to healthy controls, individuals with CAI rely on vestibular feedback to maintain posture in the injured-limb [20]. The degree of sensory system diversity or isolation may influence the regularity (readiness) and adaptability of biological noise underlying movement. For example, the greater the sensory diversity (multisensory), the more clustered sensory feedback the central nervous system (CNS) must configure to coordinate motor behaviors. Whereas the higher the sensory isolation (unisensory), the limited sensory feedback the CNS has to configure to coordinate motor behaviors. As a result, either too many or too few interactions between physiological elements (e.g., sensory feedback) affect individuals’ flexibility and adaptability to environmental constraints, resulting in either more random or rigid movement patterns.

Harborne and Stergiou [21] established a theoretical model with an inverted-U-shaped that defines optimal movement variability in connection to mature motor skills and health status. According to the theoretical model, optimal movement variability reveals the flexibility and adaptability of sensorimotor systems in healthy individuals [18]. As a healthy state, the uppermost point of the inverted-U shape represents optimal movement variability. In contrast, the points below the uppermost point of the inverted-U shape indicates inadequate movement variability associated with poor health, such as CAI, resulting in either increased or decreased optimal movement variability [21]. When optimal movement variability decreases, motor behaviors become more predictable (rigid), and when optimal variability increases, motor behaviors become unpredictable (random). Furthermore, both predictable and unpredictable motor behaviors are less flexible and adaptable to exploring environmental constraints.

According to current evidence, individuals with CAI display rigid movement patterns during walking at a self-selected speed [22]. This movement rigidity found in CAI interferes with performance, especially when environmental constraints are increased, contributing to an increased risk of recurrent ankle sprains. However, it is currently unknown how movement variability underlying postural control emerges as environmental constraints become more complex through the manipulation of somatosensory, visual, and vestibular feedback.

Therefore, the purpose of this study was to examine underlying biological noise pertaining to postural control during increased environmental constraints with sensory feedback manipulations in individuals with and without CAI.

## Methods

### Study Design

We implemented a laboratory case-control study to examine the effect of environment and task constraints on neural mechanisms underlying postural control in individuals with and without CAI.

### Participants

The study included forty-two physically active individuals with and without unilateral CAI participated in this study (Table 1). The International Ankle Consortium position statement was used to define individuals with CAI.[23] Healthy controls were matched to individuals with CAI for sex, age (years, ±2), height (cm, ±5%), mass (kg, ±3%), limb dominance (the leg used to kick a ball), and the National Aeronautics and Space Administration Physical Activity Status Scale [NASA-PASS] (scale, ±1). Healthy individuals were assigned an injured-limb based on the matched limb dominance of individuals with CAI. All participants signed the informed consent forms that were approved by the University of North Carolina Greensboro’s Institutional Review Board.

**Table 1.**
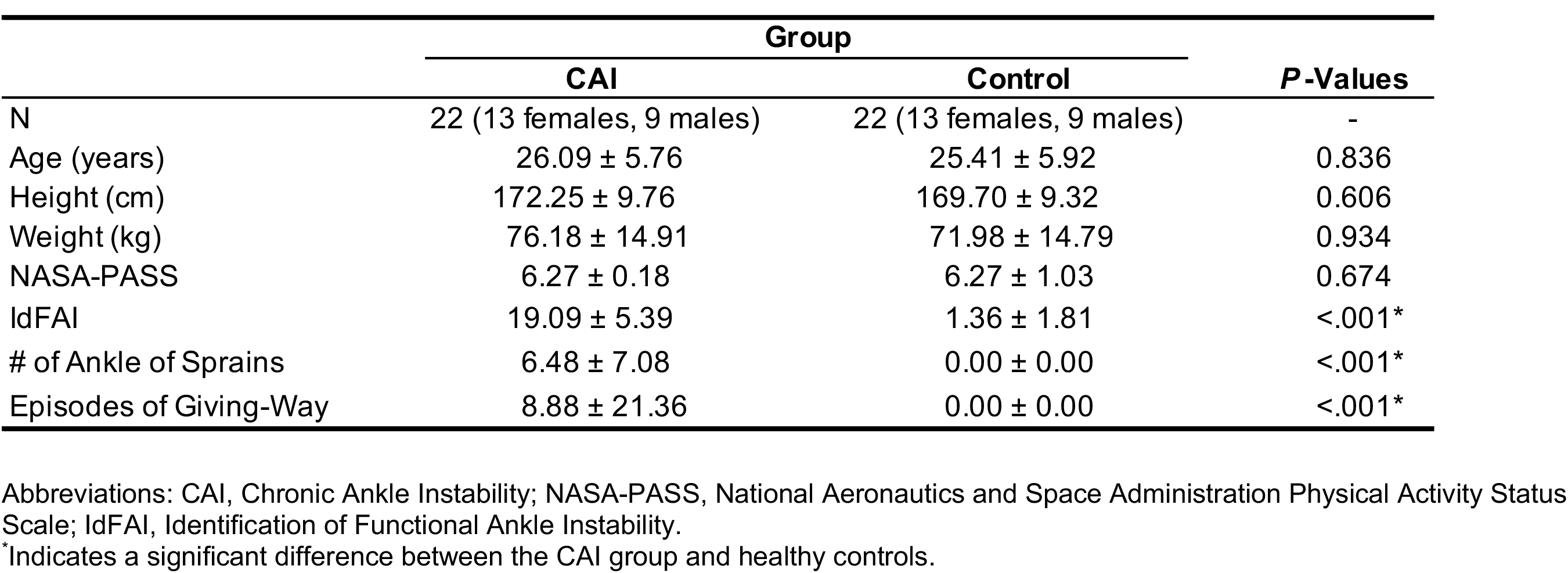
Participants’ Demographics and Patient-Reported Outcome Measure Scores (Mean±SD)

|  | Group |  | *P*-Values |
| --- | --- | --- | --- |
|  | CAI | Control |  |
| N | 22 (13 females, 9 males) | 22 (13 females, 9 males) | - |
| Age (years) | 26.09 ± 5.76 | 25.41 ± 5.92 | 0.836 |
| Height (cm) | 172.25 ± 9.76 | 169.70 ± 9.32 | 0.606 |
| Weight (kg) | 76.18 ± 14.91 | 71.98 ± 14.79 | 0.934 |
| NASA-PASS | 6.27 ± 0.18 | 6.27 ± 1.03 | 0.674 |
| IdFAI | 19.09 ± 5.39 | 1.36 ± 1.81 | <.001* |
| # of Ankle of Sprains | 6.48 ± 7.08 | 0.00 ± 0.00 | <.001* |
| Episodes of Giving-Way | 8.88 ± 21.36 | 0.00 ± 0.00 | <.001* |

Abbreviations: CAI, Chronic Ankle Instability; NASA-PASS, National Aeronautics and Space Administration Physical Activity Status Scale; IdFAI, Identification of Functional Ankle Instability.
\*Indicates a significant difference between the CAI group and healthy controls.

### Procedure

Participants with and without CAI completed a single session in a biomechanics research laboratory and a standardized medical history questionnaire including medical history of their lower extremity and completion of rehabilitation following ankle sprains, self-reported ankle instability and function, and physical activity status upon arrival. All participants warmed up for a 5-minute on a bike at a self-selected intensity and completed demographic measures (height, weight), joint hypermobility tests, lower extremity anatomical alignment measures, and postural tests in the injured-, uninjured-, and double-limbs. Hypermobility, anatomical tests, and postural control in the uninjured- and double-limbs are part of a big research study and are not reported in current results. All participants were outfitted with a vest and safety harness before being asked to stand barefoot on a computerized NeuroCom dynamic posturography platform (SMART EquiTest, NeuroCom International Inc., Clackamas, OR) in single-limb stance (Figure 1).

**Figure 1.**
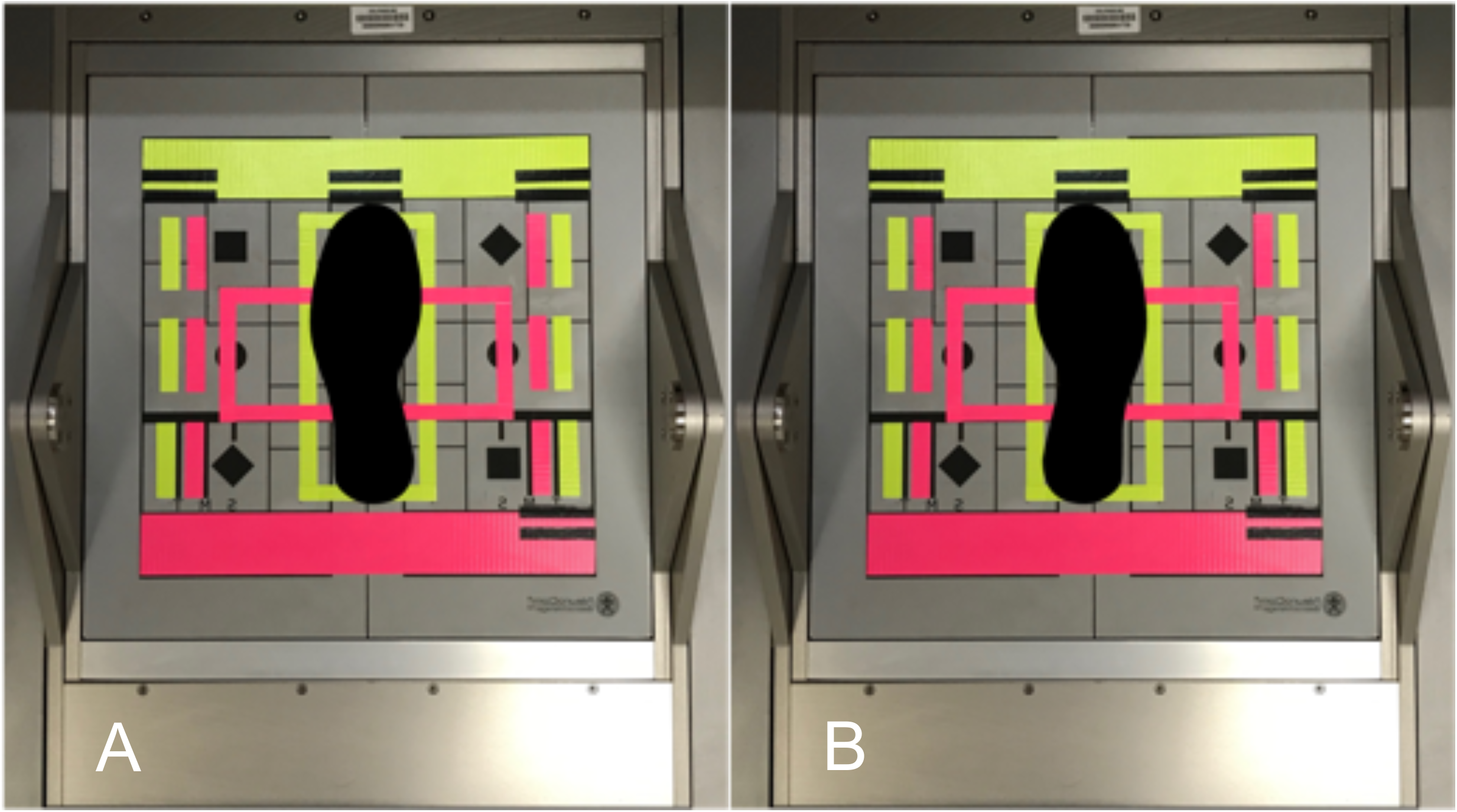
Foot Position for the Sensory Organization Test in the injured-limb (A, B)

#### Sensory Organization Test (SOT)

The SOT is a gold-standard test that examines individuals’ ability to integrate somatosensory, visual, and vestibular feedback to maintain posture. In the SOT’s six conditions (Table 2), a combination of the sway-referenced support surface (platform) and visual surroundings with and without vision is used to alter somatosensory and visual feedback. SOT conditions progress from simple to more complex environmental constraints to isolate different sensory systems as follows: 1-Normal Vision-Fixed Surroundings-Fixed Platform (C1-V_n_S_f_P_f_); 2-Absent Vision-Fixed Surroundings-Fixed Platform (C2-V_a_S_f_P_f_); 3-Distorted Vision-Moving Surroundings-Fixed Platform (C3-V_d_S_m_P_f_); 4-Distorted Vision-Fixed Surroundings-Moving Platform (C4-V_d_S_f_P_m_); 5-Absent Vision-Fixed Surroundings-Moving Platform (C5-V_a_S_f_P_m_); and 6-Distorted Vision-Moving Surroundings-Moving Platform (C6-V_d_S_m_P_m_).

**Table 2.**
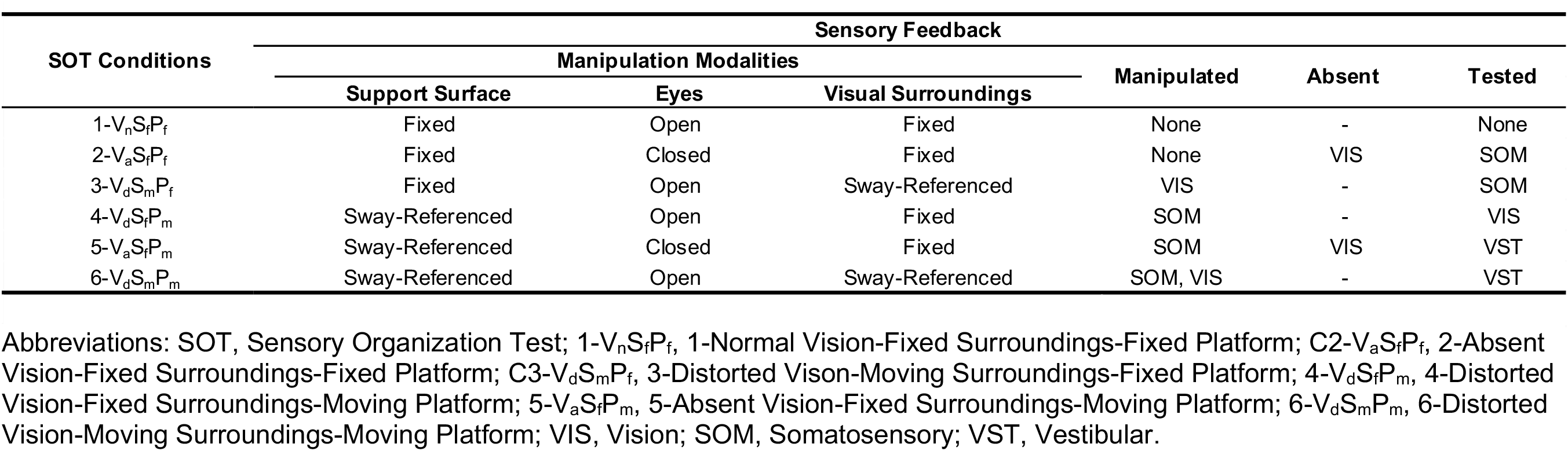
Descriptions of Six Sensory Organization Test Conditions.

Participants’ medial malleoli of the tested ankle was aligned perpendicular to the transverse axis of platform rotation, and the foot was positioned in the center of the platform for single-limb stance (Figure 1). Participants were instructed to maintain their faces forward, keep their arms relaxed by their sides, and stand motionless as possible while completing the SOT. Each condition consisted of three 20-second trials, a total of 18 trials per stance. Participants were given a 30-second rest between trials and a 1-minute rest between conditions. Each participant was allowed to tap down on the platform with non-stance toes multiple times after 10 seconds to continue with full 20-second trials in the injured-limb. To complete an individual 20-second trial, participants were instructed to make every effort to preserve single-limb postural stability. The trials were stopped, eliminated, and repeated if participants tapped down on non-stance toes before 10 seconds and/or completely stood on a non-stance limb after 10 seconds.

#### Movement Variability Measure

The SOT of the NeuroCom sampled COP coordinates for anteroposterior (AP) and mediolateral (ML) components at 100 Hz for every three trials of individual SOT conditions. The raw data from the NeuroCom were exported to spreadsheets (Excel, version 360; Microsoft Corporation, Redmond, WA) and imported into a custom R program in RStudio (version 4.0.0; RStudio, Inc., Boston, MA) to compute path length following equation 1, where *N* represents the number of data points and *i* is each successive data point [24]. Path length was calculated by summing the magnitude of the distance change of the COP at every time point from the resultant vector created from the combined COP AP and ML time based on the first 10-second trials [24].

SampEN values were computed using combination of the parameters *m* = 3 and *r* = 0.2 of the time series for COP path length with a custom R program based on an algorithm proposed by Richman and Moorman [25] [26]. The efficacy of the metric was derived by setting the maximal relative error less than 0.05, corresponding to a 95% confidence interval, which is a 10% sample entropy estimation. SampEN values typically range from 0 to 2 in human biological systems. Both lower and greater SampEN values correspond to less flexibility and adaptability of sensorimotor systems based on the theoretical model with an inverted-U-shaped presented earlier [18]. Lower SampEN values result in more predictable rigid movement patterns, whereas greater SmpEN Values result in more unpredictable random movement patterns [18], [27], [28].

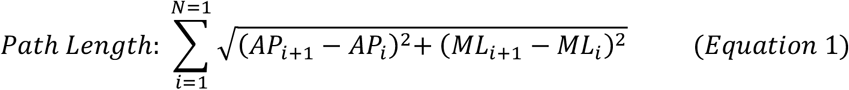

### Statistical Analysis

A separate one-way analysis of variance (ANOVA) was conducted to compare group differences in demographic characteristics (age, height, body weight, physical activity level, the number of ankle sprains, and self-reported giving-way) and Identification of Functional Ankle Instability (IdFAI). A one-way ANOVA was also used to examine group differences in biological noise underlying postural control during the SOT conditions in single-limb stance. All statistical analyses were performed using SPSS software (version 27; IBM Corp, Armonk, NY, USA) with a priori *α* level of 0.05. Data normality was tested using the Kolmogorov-Smirnov test confirming a normal distribution for all variables (*P*>.05).

## Results

No group differences were found related to age, height, weight, or physical activity level (*P*>.05; Table 1). The CAI group had a greater number of ankle sprains, self-reported giving way, and higher IdFAI scores compared to healthy controls (*P range*: <.001-.001; Table 1). Individuals with CAI demonstrated significantly lower SampEN values while maintaining posture during C5-V_a_S_f_P_m_ (*P*=.037) and C6-V_d_S_m_P_m_ (*P*=.030) in single-limb stance (Table 3). The CAI group displayed similar SampEn values while maintaining posture during C1-V_n_S_f_P_f_, C2-V_a_S_f_P_f_, C3-V_d_S_m_P_f_, and C4-V_d_S_f_P_m_ (*P*>.05; Table 3).

**Table 3.**
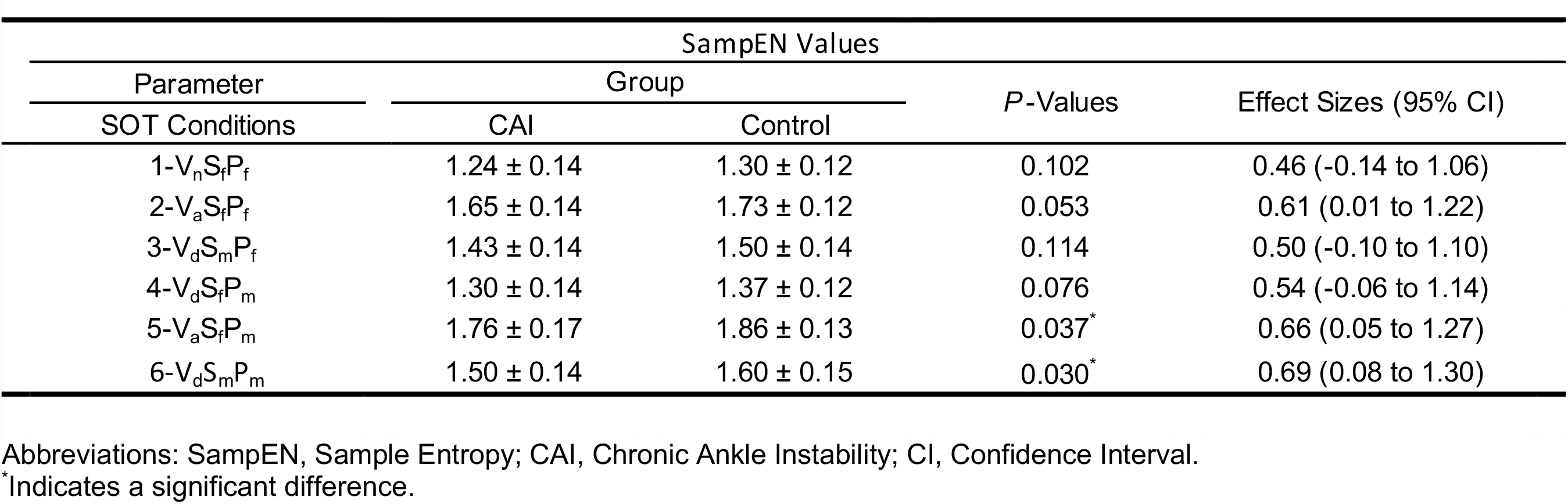
Group Differences for Movement Variability in the Injured-Limb (Mean±SD)

| Parameter<br>SOT Conditions | SampEN Values |  |  |  |
| --- | --- | --- | --- | --- |
|  | Group |  | P-Values | Effect Sizes (95% CI) |
|  | CAI | Control |  |  |
| 1-V<sub>n</sub>S<sub>f</sub>P<sub>f</sub> | 1.24 ± 0.14 | 1.30 ± 0.12 | 0.102 | 0.46 (-0.14 to 1.06) |
| 2-V<sub>a</sub>S<sub>f</sub>P<sub>f</sub> | 1.65 ± 0.14 | 1.73 ± 0.12 | 0.053 | 0.61 (0.01 to 1.22) |
| 3-V<sub>d</sub>S<sub>m</sub>P<sub>f</sub> | 1.43 ± 0.14 | 1.50 ± 0.14 | 0.114 | 0.50 (-0.10 to 1.10) |
| 4-V<sub>d</sub>S<sub>f</sub>P<sub>m</sub> | 1.30 ± 0.14 | 1.37 ± 0.12 | 0.076 | 0.54 (-0.06 to 1.14) |
| 5-V<sub>a</sub>S<sub>f</sub>P<sub>m</sub> | 1.76 ± 0.17 | 1.86 ± 0.13 | 0.037* | 0.66 (0.05 to 1.27) |
| 6-V<sub>d</sub>S<sub>m</sub>P<sub>m</sub> | 1.50 ± 0.14 | 1.60 ± 0.15 | 0.030* | 0.69 (0.08 to 1.30) |

Abbreviations: SampEN, Sample Entropy; CAI, Chronic Ankle Instability; CI, Confidence Interval.
\*Indicates a significant difference.

## Discussion

The current study is the first to investigate how the underlying biological noise in unilateral postural control changes with increased environmental constraints with sensory feedback manipulation in CAI. The primary findings were that individuals with CAI displayed lower movement variability in postural control during C5-V_a_S_f_P_m_ with the absent vision-fixed surroundings-moving platform and C6-V_d_S_m_P_m_ with the distorted vision-moving surroundings-moving platform in single-limb stance compared to healthy controls. The CAI group also displayed a moderate trend (effect size=0.61 with 95% confidence interval not crossing zero) of lower movement variability in postural control during C2-V_a_S_f_P_f_ with the absent vision-fixed surroundings-fixed platform.

Harborne and Stergiou [21] established a theoretical model of movement variability with an inverted-U-shaped as it relates to mature motor skills and health state. According to researchers, the theoretical model characterizes optimal movement variability as the flexibility and adaptability of sensorimotor systems in healthy individuals [18]. In particular, the uppermost point of the inverted-U shape represents optimal movement variability as a healthy state. In contrast, the lower or higher points below the uppermost point of the inverted-U shape suggest inadequate variability linked with poor health [21]. The development of CAI is thought to impair the healthy state of optimal variability, resulting in lower SampEN values during walking in individuals with CAI compared to healthy controls [22]. In the case of postural control, a decrease in optimal movement variability leads to more predictable (rigid) COP excursion, while increasing optimal variability leads to unpredictable (random) COP excursion.

According to current literature, individuals with CAI displayed decreased movement variability of stride-to-stride frontal plane ankle kinematics compared to healthy controls [22]. Moreover, lower movement variability relevant to postural control and gait mechanics has also been described in individuals with neurological deficits that are not confined to CAI [e.g., proprioceptive impairments, concussion, anterior cruciate ligament damage] [29]–[32]. Although our study findings did not display lower movement variability throughout the SOT conditions, lower movement variability was exhibited during C5-V_a_S_f_P_m_ and C6-V_d_S_m_P_m_ compared to healthy controls.

Environmental constraints become difficult as the SOT conditions evolve (table?) with different combinations of sensory feedback manipulations. The C5-V_a_S_f_P_m_ and C6-V_d_S_m_P_m_ are the SOT conditions where participants were forced to exclusively rely on vestibular feedback when somatosensory and visual feedback became simultaneously unreliable. To maintain postural stability, healthy individuals rely on multisensory feedback from the somatosensory, visual, and vestibular systems. The alterations of these primary sensory feedback result in the central nervous system placing emphasis on the most relevant sensory feedback to maintain postural stability. Therefore, less flexible, and adaptable sensorimotor systems associated with CAI become the most prominent when environmental constraints become more complex and individuals with CAI are forced to rely on vestibular feedback, resulting in rigid motor behavior while maintaining posture in the injured-limb during the C5-V_a_S_f_P_m_ and C6-V_d_S_m_P_m_.

Interestingly, there was a moderate trend of lower movement variability in postural control during C2-V_a_S_f_P_f_ where participants are forced to rely on somatosensory feedback with the absent vision-fixed surroundings-fixed platform. The SOT conditions are designed to manipulate somatosensory and visual feedback in a combination of the sway-referenced support surface and visual surroundings with and without absent vision. Current evidence suggests closing the eyes result in the deactivation of the visual cortex [33]. Therefore, vestibular feedback becomes the primary sensory system to facilitate somatosensory sensitivity when vision become absent [34]. This study was part of a larger experiment, and our CAI participants did not display somatosensory dysfunction and maintained postural control in the injured-limb compatible with healthy controls (Sugimoto et al., 2023). Consequently, our participants with CAI displayed lower movement variability when vestibular feedback became primarily sensory feedback to maintain postural control in the injured-limb. Indeed, the CAI group particularly failed to downregulate vestibular feedback reliance while maintaining posture in the injured-limb than healthy controls (Sugimoto et al., 2023).

The study’s limitation of the research was that we had individuals who had participated in a variety of sports, not just collegiate sports. In addition, individuals with CAI in our study reported having finished some form of rehabilitation following an initial ankle sprain. As a consequence, the findings may not be applicable to individuals with diverse sports participation histories or those who did not finish any rehabilitation following an initial ankle sprain, whether within or outside of the recruited age group.

## Conclusion

Although decreased or increased movement variability is associated with poor health, such as CAI, our CAI participants did not show decreased movement variability while maintaining posture in the injured-limb during all six SOT conditions when compared to healthy controls. However, when the CAI group was forced to rely exclusively on vestibular feedback to maintain postural control in the injured-limb, they demonstrated decreased movement variability. Future studies should investigate how vestibular feedback manipulation impacts movement variability underlying postural control in individuals with CAI.

## Data Availability

All data produced in the present work are contained in the manuscript

## Declaration of Interest

None

## Ethical Approval

All participants signed the informed consent forms that were approved by the University of North Carolina Greensboro’s Institutional Review Board.

## Notes

**Funding:** Research reported in this publication was supported by the NATA Research and Education Foundation Doctoral Grant.

**Conflicts of Interest:** None

### Competing Interest Statement

The authors have declared no competing interest.

### Author Declarations

IRB of the University of North Carolina at Greensboro gave ethical approval for this work.

## References

[1] J. M. Medina McKeon and M. C. Hoch, “The Ankle-Joint Complex: A Kinesiologic Approach to Lateral Ankle Sprains,” J. Athl. Train., vol. 54, no. 6, pp. 589–602, Jun. 2019, doi: 10.4085/1062-6050-472-17.

[2] P. Kannus and P. Renström, “Treatment for acute tears of the lateral ligaments of the ankle. Operation, cast, or early controlled mobilization.:,” J. Bone Jt. Surg., vol. 73, no. 2, pp. 305–312, Feb. 1991, doi: 10.2106/00004623-199173020-00021.

[3] B. R. Waterman, P. J. Belmont, K. L. Cameron, T. M. DeBerardino, and B. D. Owens, “Epidemiology of Ankle Sprain at the United States Military Academy,” Am. J. Sports Med., vol. 38, no. 4, pp. 797–803, Apr. 2010, doi: 10.1177/0363546509350757.

[4] E. A. Wikstrom, T. Hubbard-Turner, S. Guderian, and M. J. Turner, “Lateral Ankle Sprain in a Mouse Model: Lifelong Sensorimotor Dysfunction,” J. Athl. Train., vol. 53, no. 3, pp. 249–254, Mar. 2018, doi: 10.4085/1062-6050-365-16.

[5] A. Anandacoomarasamy, L. Barnsley, and L. Grujic, “Long term outcomes of inversion ankle injuries,” Br. J. Sports Med., vol. 39, no. 3, p. e14, Mar. 2005, doi: 10.1136/bjsm.2004.011676.

[6] A. C. Thomas, T. Hubbard-Turner, E. A. Wikstrom, and R. M. Palmieri-Smith, “Epidemiology of Posttraumatic Osteoarthritis,” J. Athl. Train., vol. 52, no. 6, pp. 491–496, Jun. 2017, doi: 10.4085/1062-6050-51.5.08.

[7] B. L. Arnold, C. J. Wright, and S. E. Ross, “Functional Ankle Instability and Health-Related Quality of Life,” J. Athl. Train., vol. 46, no. 6, pp. 634–641, 2011.

[8] S. A. Hale, J. Hertel, and L. C. Olmsted-Kramer, “The Effect of a 4-Week Comprehensive Rehabilitation Program on Postural Control and Lower Extremity Function in Individuals With Chronic Ankle Instability,” J. Orthop. Sports Phys. Ther., vol. 37, no. 6, pp. 303–311, Jun. 2007, doi: 10.2519/jospt.2007.2322.

[9] T. J. Hubbard, L. C. Kramer, C. R. Denegar, and J. Hertel, “Contributing Factors to Chronic Ankle Instability,” Foot Ankle Int., vol. 28, no. 3, pp. 343–354, Mar. 2007, doi: 10.3113/FAI.2007.0343.

[10] A. J. Y. Lee, W.-H. Lin, and C. H. Huang, “IMPAIRED PROPRIOCEPTION AND POOR STATIC POSTURAL CONTROL IN SUBJECTS WITH FUNCTIONAL INSTABILITY OF THE ANKLE,” J Exerc Sci Fit, vol. 4, no. 2, pp. 117–125, 2006.

[11] S. E. Ross and K. M. Guskiewicz, “Examination of Static and Dynamic Postural Stability in Individuals With Functionally Stable and Unstable Ankles:,” Clin. J. Sport Med., vol. 14, no. 6, pp. 332–338, Nov. 2004, doi: 10.1097/00042752-200411000-00002.

[12] H. Tropp, J. Ekstrand, and J. Gillquist, “Factors affecting stabilometry recordings of single limb stance,” Am. J. Sports Med., vol. 12, no. 3, pp. 185–188, May 1984, doi: 10.1177/036354658401200302.

[13] X. Xue et al., “Postural Control Deficits During Static Single-leg Stance in Chronic Ankle Instability: A Systematic Review and Meta-Analysis,” Sports Health Multidiscip. Approach, p. 194173812311524, Mar. 2023, doi: 10.1177/19417381231152490.

[14] P. O. Mckeon, C. D. Ingersoll, D. C. Kerrigan, E. Saliba, B. C. Bennett, and J. Hertel, “Balance Training Improves Function and Postural Control in Those with Chronic Ankle Instability:,” Med. Sci. Sports Exerc., vol. 40, no. 10, pp. 1810–1819, Oct. 2008, doi: 10.1249/MSS.0b013e31817e0f92.

[15] S. M. Glass, S. E. Ross, B. L. Arnold, and C. K. Rhea, “Center of Pressure Regularity With and Without Stochastic Resonance Stimulation in Stable and Unstable Ankles,” Athl. Train. Sports Health Care, vol. 6, no. 4, pp. 170–178, Jul. 2014, doi: 10.3928/19425864-20140710-04.

[16] C. P. Hoffmann, B. Seigle, J. Frère, and C. Parietti-Winkler, “Dynamical analysis of balance in vestibular schwannoma patients,” Gait Posture, vol. 54, pp. 236–241, May 2017, doi: 10.1016/j.gaitpost.2017.03.015.

[17] I.-C. Lee, M. M. Pacheco, and K. M. Newell, “A test of fixed and moving reference point control in posture,” Gait Posture, vol. 51, pp. 52–57, Jan. 2017, doi: 10.1016/j.gaitpost.2016.09.028.

[18] N. Stergiou and L. M. Decker, “Human movement variability, nonlinear dynamics, and pathology: Is there a connection?,” Hum. Mov. Sci., vol. 30, no. 5, pp. 869–888, Oct. 2011, doi: 10.1016/j.humov.2011.06.002.

[19] J. Hertel and R. O. Corbett, “An Updated Model of Chronic Ankle Instability,” J. Athl. Train., vol. 54, no. 6, pp. 572–588, Jun. 2019, doi: 10.4085/1062-6050-344-18.

[20] Y. A. Sugimoto, C. K. Rhea, and S. E. Ross, “Modified proximal thigh kinematics captured with a novel smartphone app in individuals with a history of recurrent ankle sprains and altered dorsiflexion with walking,” Clin. Biomech., vol. 105, p. 105955, May 2023, doi: 10.1016/j.clinbiomech.2023.105955.

[21] R. T. Harbourne and N. Stergiou, “Movement Variability and the Use of Nonlinear Tools: Principles to Guide Physical Therapist Practice,” Phys. Ther., vol. 89, no. 3, pp. 267–282, Mar. 2009, doi: 10.2522/ptj.20080130.

[22] M. Terada et al., “Alterations in stride-to-stride variability during walking in individuals with chronic ankle instability,” Hum. Mov. Sci., vol. 40, pp. 154–162, Apr. 2015, doi: 10.1016/j.humov.2014.12.004.

[23] P. A. Gribble et al., “Selection Criteria for Patients With Chronic Ankle Instability in Controlled Research: A Position Statement of the International Ankle Consortium,” J. Athl. Train., vol. 49, no. 1, pp. 121–127, Jan. 2014, doi: 10.4085/1062-6050-49.1.14.

[24] C. K. Rhea, A. W. Kiefer, F. J. Haran, S. M. Glass, and W. H. Warren, “A new measure of the CoP trajectory in postural sway: Dynamics of heading change,” Med. Eng. Phys., vol. 36, no. 11, pp. 1473–1479, Nov. 2014, doi: 10.1016/j.medengphy.2014.07.021.

[25] J. S. Richman and J. R. Moorman, “Physiological time-series analysis using approximate entropy and sample entropy,” Am. J. Physiol.-Heart Circ. Physiol., vol. 278, no. 6, pp. H2039–H2049, Jun. 2000, doi: 10.1152/ajpheart.2000.278.6.H2039.

[26] D. E. Lake, J. S. Richman, M. P. Griffin, and J. R. Moorman, “Sample entropy analysis of neonatal heart rate variability,” Am. J. Physiol.-Regul. Integr. Comp. Physiol., vol. 283, no. 3, pp. R789–R797, Sep. 2002, doi: 10.1152/ajpregu.00069.2002.

[27] J. M. Yentes, N. Hunt, K. K. Schmid, J. P. Kaipust, D. Mcgrath, and N. Stergiou, “The Appropriate Use of Approximate Entropy and Sample Entropy with Short Data Sets,” Ann. Biomed. Eng. N. Y., vol. 41, no. 2, pp. 349–65, Feb. 2013, doi: 10.1007/s10439-012-0668-3.

[28] J. M. Yentes, W. Denton, J. McCamley, P. C. Raffalt, and K. K. Schmid, “Effect of parameter selection on entropy calculation for long walking trials,” Gait Posture, vol. 60, pp. 128–134, Feb. 2018, doi: 10.1016/j.gaitpost.2017.11.023.

[29] J. T. Cavanaugh, “Detecting altered postural control after cerebral concussion in athletes with normal postural stability,” Br. J. Sports Med., vol. 39, no. 11, pp. 805–811, Nov. 2005, doi: 10.1136/bjsm.2004.015909.

[30] B. Manor et al., “Altered control of postural sway following cerebral infarction,” Neurology, vol. 74, no. 6, pp. 458–464, Feb. 2010, doi: 10.1212/WNL.0b013e3181cef647.

[31] C. Moraiti, N. Stergiou, S. Ristanis, and A. D. Georgoulis, “ACL deficiency affects stride-to-stride variability as measured using nonlinear methodology,” Knee Surg. Sports Traumatol. Arthrosc., vol. 15, no. 12, pp. 1406–1413, Dec. 2007, doi: 10.1007/s00167-007-0373-1.

[32] J. J. Sosnoff et al., “Mobility, Balance and Falls in Persons with Multiple Sclerosis,” PLoS ONE, vol. 6, no. 11, Nov. 2011, doi: 10.1371/journal.pone.0028021.

[33] K. Uludağ, D. J. Dubowitz, E. J. Yoder, K. Restom, T. T. Liu, and R. B. Buxton, “Coupling of cerebral blood flow and oxygen consumption during physiological activation and deactivation measured with fMRI,” NeuroImage, vol. 23, no. 1, pp. 148–155, Sep. 2004, doi: 10.1016/j.neuroimage.2004.05.013.

[34] C. Pfeiffer, J. Noel, A. Serino, and O. Blanke, “Vestibular modulation of peripersonal space boundaries,” Eur. J. Neurosci., vol. 47, no. 7, pp. 800–811, Apr. 2018, doi: 10.1111/ejn.13872.

